# The ENGAGE Study: A Randomized Trial Optimizing Uptake of Germline Cancer Genetic Services in Childhood Cancer Survivors

**DOI:** 10.1101/2025.10.20.25338173

**Authors:** Tara O. Henderson, Brian Egleston, Sarah Howe, Mary Ashley Allen, Rajia Mim, Linda G. Fleisher, Elena B. Elkin, Kevin C. Oeffinger, Kevin R. Krull, Demetrios Ofidis, Briana Mcleod, Hannah Griffin, Elisabeth M. Wood, Cara N. Cacioppo, Sarah Brown, Melody Perpich, Gregory T. Armstrong, Angela R. Bradbury

## Abstract

**Background:** Identifying childhood cancer survivors who are already at high risk of subsequent neoplasms and may also have an inherited genetic susceptibility is essential for effective surveillance and prevention. This trial evaluated the effectiveness of remote, centralized telehealth genetic services in increasing service uptake.

**Methods:** Childhood Cancer Survivor Study (CCSS) participants at the St. Jude Research Hospital, who were ≥18 years old and survivors of a CNS tumor, sarcoma, or more than one primary cancer, were recruited for the study. After completing a baseline survey, participants were randomly assigned to one of three arms: remote telehealth genetic services via phone, videoconference, or usual care. Uptake of genetic services was obtained through study records and the six-month Status Survey.

**Findings:** Of the 391 participants recruited, 262 were assigned to remote telehealth services (via phone or videoconference) and 129 to usual care. At six months, 43% (113/262) of participants in remote telehealth services received genetic services compared to 15% (19/129) in the usual care group (OR = 4·4, 95% CI 2·5-8·0, p<0·0001). Uptake of genetic counseling (42% vs. 15%, p < 0·0001) and genetic testing (19% vs. 9%, p = 0·020) were higher in remote telehealth services. Factors associated with higher uptake included not having high-deductible health insurance (OR = 1·67, 95%CI 1·00-2·91, p = 0·049) and lower perceived cost of testing (OR = 1·51, 95%CI 1·17-1·96, p = 0·0014). Top barriers included experiencing higher levels of depression (OR=0·91, 95%CI 0·85-0·98, p = 0·0067) and anxiety (OR=0·93, 95%CI 0·87-1·00, p = 0·036).

**Interpretation:** Remote telehealth genetic services improve genetic counseling and testing uptake in childhood cancer. Addressing remaining barriers could maximize their impact and ensure equitable access for childhood cancer survivors and their families.

**Funding:** National Cancer Institute (R01-CA237369, U24-CA55727).

**Clinical Trial Registration:** NCT 04455698 at http://www.ClinicalTrials.gov

## INTRODUCTION

The five-year survival rate for childhood cancer has exceeded 85%, resulting in a growing population of childhood cancer survivors in the United States.^1^ These survivors face a lifelong risk of premature mortality.^2^ Subsequent malignant neoplasms (SMNs) are the leading cause of non-relapse-related premature mortality in this population.^3^ While SMNs are primarily linked to prior cancer therapies,^4^ inherited genetic susceptibilities can also increase the risk in both irradiated and non-irradiated survivors.^5,6^ Recent studies have reported that 10% to 13% of childhood cancer survivors carry *moderate- and high-penetrance dominant pathogenic/likely pathogenic (P/LP) variants*.^5–7^ Survivors with such mutations have a fourfold increased risk of early mortality due to SMNs.^8^

The National Academy of Medicine recommends lifelong, risk-based healthcare for childhood cancer survivors,^8^ prompting the Children’s Oncology Group (COG) to develop exposure-based, long-term follow-up guidelines, including genetic risk assessment.^9^ Identifying survivors with germline mutations enables personalized cancer preventive strategies, such as lifestyle changes, enhanced screening, chemoprevention, or prophylactic surgery.^5,7^ However, over 85% of survivors rely on primary care providers (PCPs) in community settings, where access to genetic services is limited.^10^ While PCPs report a willingness to care for these survivors, many are unfamiliar with survivorship care guidelines.^11^ Even when aware, geographic disparities and workforce shortages frequently hinder access to genetic services.^12^ As a result, most survivors with germline cancer susceptibility remain undiagnosed, increasing their risk for late-stage SMNs and poor outcomes.

In a previous study among adults with cancer, offering remote telehealth genetic services via phone or videoconference significantly increased the uptake of genetic services in community medical practices (80% vs. 16% in usual care), with patient-reported outcomes comparable to in-person services.^13^ Building on this, the ENGAGE study (NCT 04455698) was conducted to utilize centralized telehealth approaches to enhance access to genetic services and adherence to guideline-recommended evaluations among childhood cancer survivors. This randomized controlled trial compared an in-home, collaborative PCP model of remote telehealth genetic service delivery with usual care, which involved accessing genetic services locally. The primary objective was to assess whether the remote centralized telehealth delivery model improved uptake of genetic services (pre-test counseling and/or genetic testing) six months after enrollment. A secondary objective was to identify factors that facilitated or hindered the uptake of genetic services overall and within the remote centralized telehealth arm.

## METHODS

### Participants

For this institutional review board approved study, participants were recruited from the Childhood Cancer Survivor Study (CCSS). CCSS is a 31-institution retrospective cohort study that comprises 25,735 long-term childhood cancer survivors who were diagnosed before age 21 between 1970 and 1999 and resided in the U.S. and Canada, as described elsewhere.^14^ In this study, we specifically targeted childhood cancer survivors with a history of central nervous system (CNS) tumors and sarcoma, given the established evidence linking these malignancies to cancer predisposition syndromes.^15,16^ Within the CCSS cohort, 7,621/25,735 (30%) had a primary diagnosis of CNS tumor or sarcoma (excluding Ewing sarcoma). Survivors were eligible for the ENGAGE study if they were 18 years or older, able to understand and communicate in English, resided in the U.S., had a history of a CNS tumor, sarcoma (excluding Ewing sarcoma), or more than one primary cancer, and had not had prior genetic testing, which was asked during screening. All participants were provided with information and a video on the benefits of genetic testing.

### Random Assignment and Study Interventions

Participants were recruited by the CCSS Coordinating Center between August 2021 and July 2023. After completing informed consent and a baseline survey, participants were randomly assigned to one of three arms: usual care or telehealth genetic services (via phone or videoconference) using a permuted block design (alternating block sizes of 3 and 6) and stratified by gender. The primary aim of the ENGAGE study was to compare the two remote telehealth service arms together with local usual care options, focusing on establishing the value of remote services compared to usual care options for testing. Comparison of longitudinal patient outcomes between the two telehealth arms was a secondary aim and will, therefore, be reported in a future publication.

In both the remote and usual care arms, participants are informed about the value of genetic testing and provided with a flyer outlining the next steps. Participants initiate services by following the instructions provided in the study flyer specific to their arm. Details of the study protocol have been published previously.^17^

Participants in the telehealth genetic services arms received an informational flyer describing how to contact the Penn Telegenetics Program to schedule an appointment.^18^ The videoconference group was provided a secure link to download BlueJeans, a HIPAA-compliant videoconferencing platform. Genetic counselors (GCs) at the Penn Telegenetics Program used standardized communication protocols, visual aids, and counseling checklists. GCs worked with participants to select appropriate tests and laboratories, such as Invitae and Ambry Genetics. Test kits were mailed directly to participants’ homes, with genetic testing costs billed to insurance consistent with real-world practice. Consistent with clinical practice, GCs worked with the participant to identify the lowest possible cost and financial assistance programs if insurance did not cover testing. Based on the in-home collaborative PCP model, a local provider was required to register with the Penn Telegenetics Program and be included in the test order. Genetic test results were shared with participants, and their local providers were provided with a summary note that included the test results and their implications.

Participants in the usual care arm received a flyer outlining local options for accessing genetic services. These included using the National Society of Genetic Counselors’ website to find a genetic counselor near them or calling the National Cancer Institute information line (1-800-4-CANCER) for local resources. They were also instructed to consult with their primary care providers or other providers for local genetic services. Research staff contacted participants in all arms to confirm receipt of the flyer and ensure comprehension.

### Assessment of Study Outcomes

The primary outcome was uptake of genetic services, defined as a composite variable indicating whether a participant had pre-test counseling and/or genetic testing within six months after enrollment. This definition accounts for participants who declined testing based on an informed decision during this timeframe. The uptake of genetic services was obtained through GC records for the telehealth genetic services arms, the 6-month post-enrollment survey for the usual care arm, and participants in the telehealth genetic services arms who lacked evidence of completing genetic counseling in service records. The 6-month post-enrollment survey included two questions for the primary outcome. “*Have you completed genetic counseling/testing in the past six months?*”. Participants were asked open and closed-ended questions about how they obtained genetic services (provider, setting, method, and cost) and why they did not complete counseling or testing, if applicable.

Measures to assess factors associated with the uptake of genetic services were collected at the baseline. These included patient-reported outcomes described below, sociodemographic characteristics, educational attainment, health insurance coverage (including whether participants had a high deductible plan, defined as >$1400), residential setting (urban/suburban/rural as determined by zip code using Federal Department of Education definitions),^19^ internet access, and health literacy.^20^

- *Knowledge of genetic disease* was evaluated using the KnowGene Scale,^21^ a 16-item tool to assess understanding of the health implications of cancer genetic testing results (Cronbach’s α 0·74 – 0·85).
- *Perception of genetic disease* was assessed with two items evaluating the perceived risk of developing cancer again on a five-point verbal scale and numerical risk (0-100%).^22^
- *General anxiety and depression* were measured using the four-item short form of the Patient Reported Outcomes Measurement Information System (PROMIS) (Cronbach’s α 0·82 – 0·91 for anxiety, Cronbach’s α 0·88 – 0·93 for depression).^23^
- *Disease-specific distress* was assessed using the eight-item Impact of Events Scale (IES) in genetic delivery studies (Cronbach’s α 0·87 – 0·95).^24^

### Statistical Analysis

The primary comparison was between usual care and telehealth genetic services, as assessed by a Fisher’s Exact test. The target accrual was 120 participants in the usual care arm and 240 in the telehealth genetic services via phone or video conference. This provided >99% power to detect a difference in the testing of 17% (usual care arm) versus 53% (telehealth arm) with a 5% Type I error rate (2-sided). In secondary analyses, we used exact logistic regressions to investigate factors associated with uptake; unadjusted odds ratios (ORs) are presented in the results, and multivariable models are in the supplement. STATAv18 (College Station, TX) was used for the analyses.

Furthermore, investigators with experience in qualitative research reviewed a subsample of open-ended responses from the six-month post-enrollment survey and developed a thematic framework of primary and secondary themes for each item (AB, BM). Two research staff with experience in qualitative methods (BM, MA) independently assigned thematic codes to all open-ended responses. Differences in code assignments were resolved through discussion (BM, MA, AB) and the establishment of an agreement for all responses.

## RESULTS

Of the 558 CCSS participants residing in the US who met the inclusion criteria and were successfully contacted, 74% (414/558) consented. Ninety-four percent (391/414) completed the baseline survey and were randomized: 262 to the remote telehealth service arms (phone or videoconference) and 129 to the usual care arm (Figure 1). Eligible non-participants (n =144) did not differ significantly from participants who consented (Supplementary Table 1).

**Figure 1.**
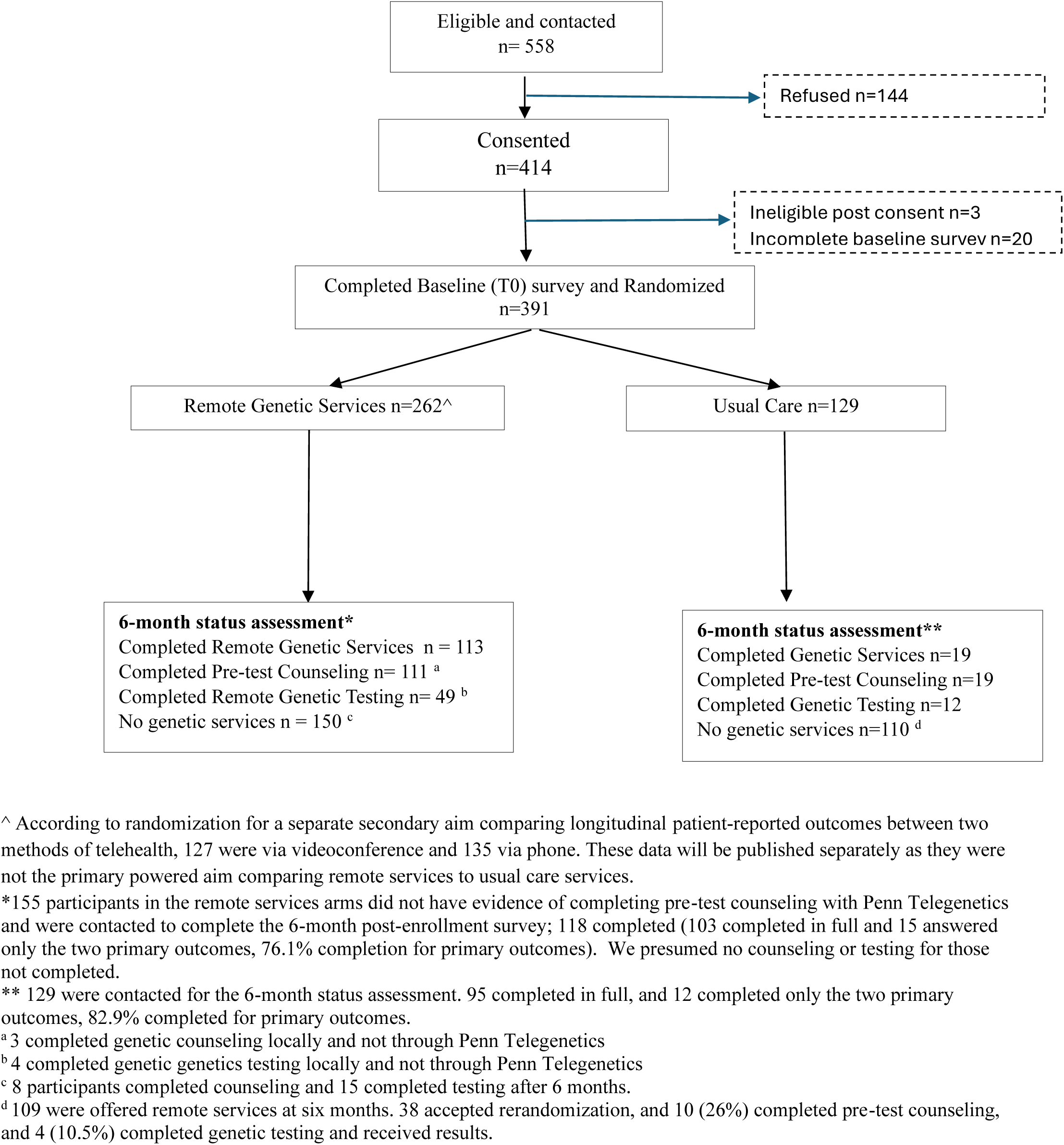
Consort Diagram.

The mean age of the participants was 44·0 (standard deviation [SD] 9·7). Twenty percent (78 / 391) identified as racial minorities, and 36% (142/391) had less than a college degree. Participants represented 40 U.S. states (Figure 2), with 47% (182/391) living in rural areas. Seventy-eight percent (204/262) of participants in the telehealth arms reported having at least one first-degree and/or second-degree relative with cancer, compared to 80% (103/129) in the usual care arm. Most participants had health insurance 95%372/391), but 34% (132/391) reported a high-deductible plan with a deductible of at least $1,400 per year (Table 1).

**Figure 2.**
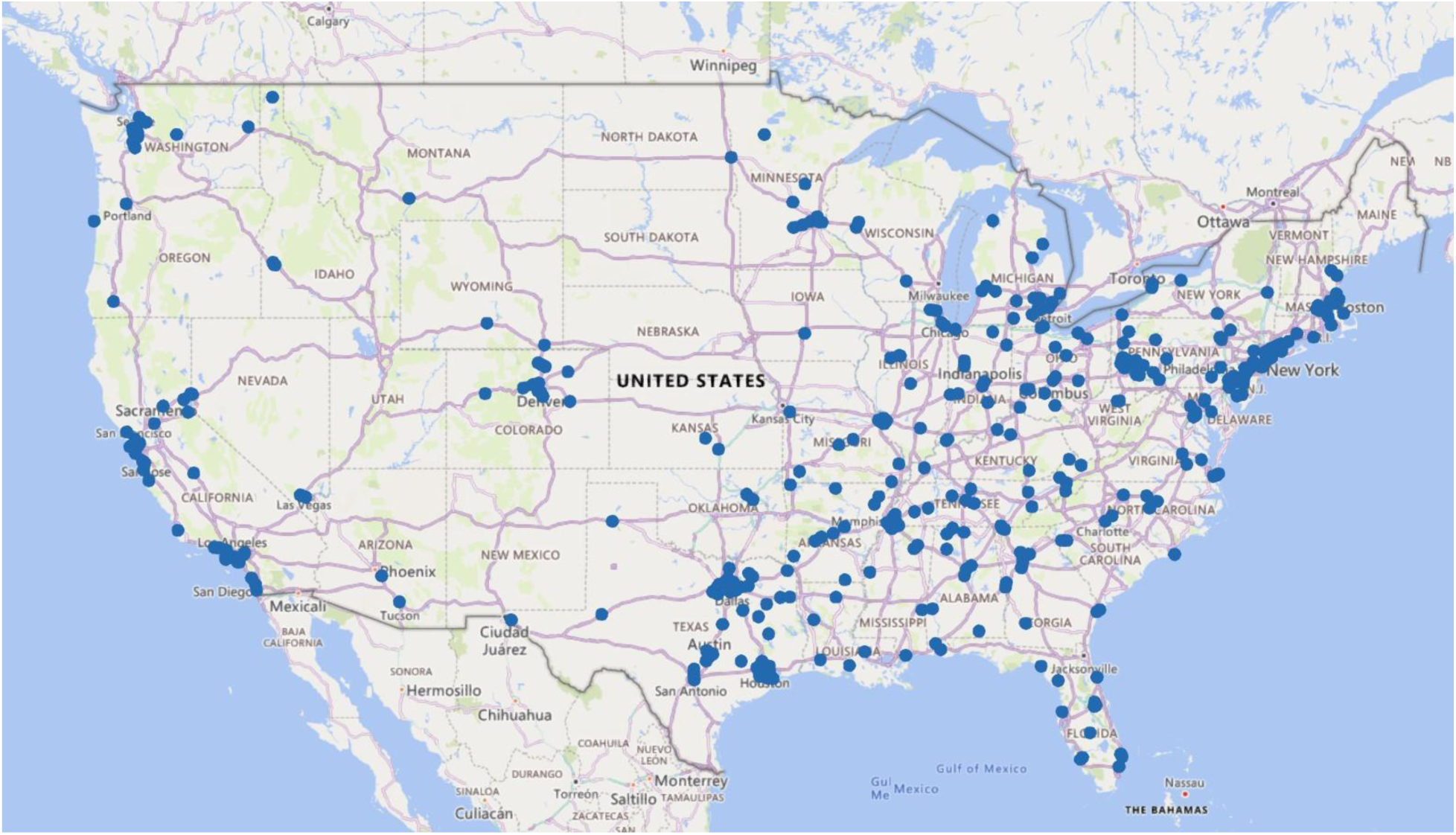
Map of participant locations.

**Table 1.**
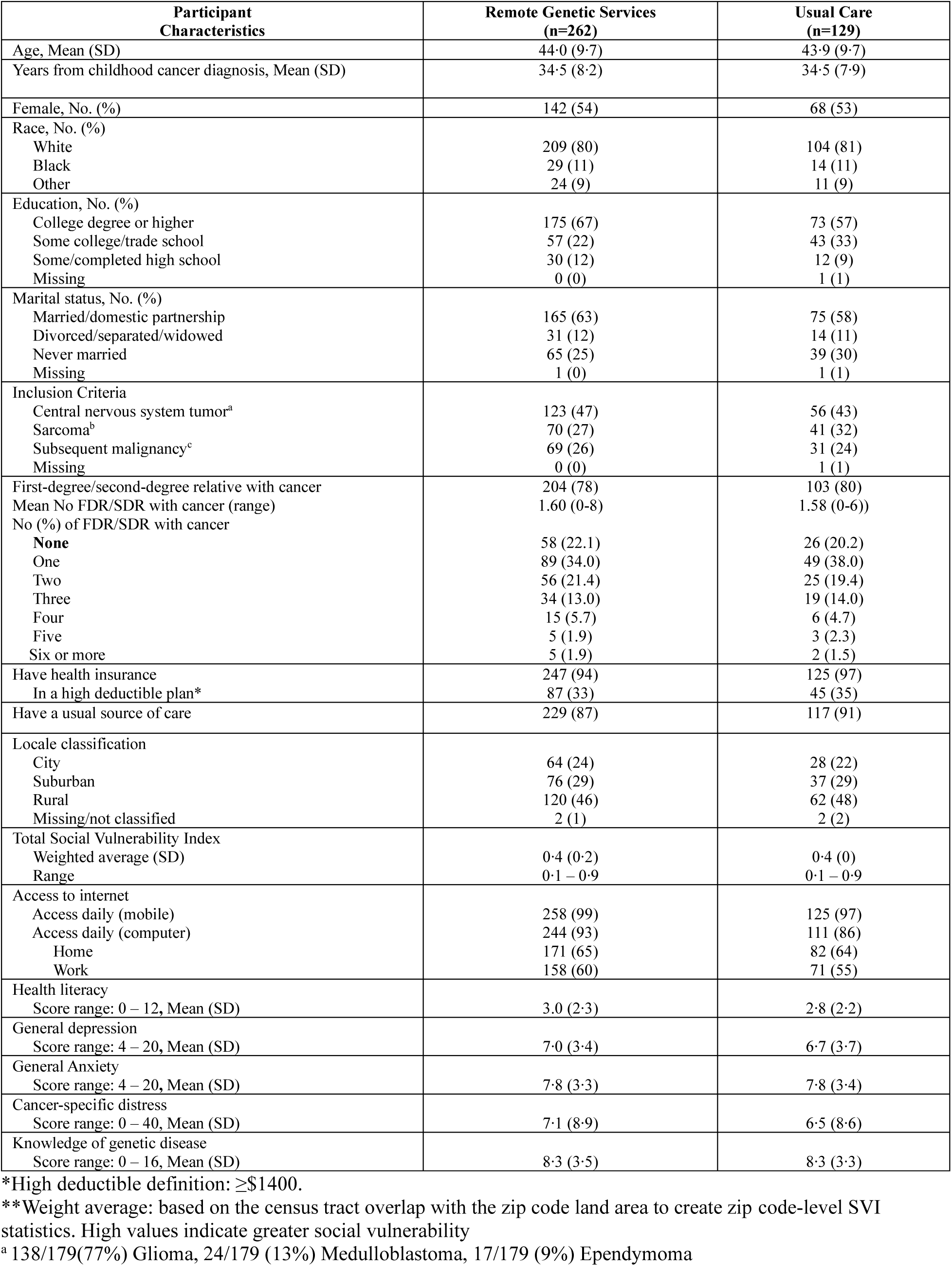
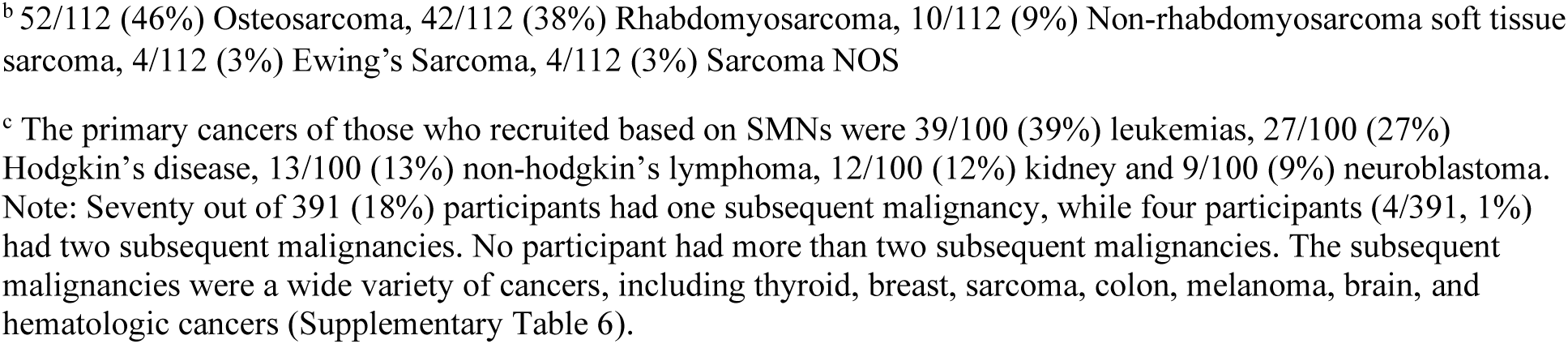
Demographic Characteristics of Enrolled Childhood Cancer Survivors (N=391).

| **Participant Characteristics** | **Remote Genetic Services (n=262)** | **Usual Care (n=129)** |
| --- | --- | --- |
| Age, Mean (SD) | 44.0 (9.7) | 43.9 (9.7) |
| Years from childhood cancer diagnosis, Mean (SD) | 34.5 (8.2) | 34.5 (7.9) |
| Female, No. (%) | 142 (54) | 68 (53) |
| Race, No. (%) |  |  |
| White | 209 (80) | 104 (81) |
| Black | 29 (11) | 14 (11) |
| Other | 24 (9) | 11 (9) |
| Education, No. (%) |  |  |
| College degree or higher | 175 (67) | 73 (57) |
| Some college/trade school | 57 (22) | 43 (33) |
| Some/completed high school | 30 (12) | 12 (9) |
| Missing | 0 (0) | 1 (1) |
| Marital status, No. (%) |  |  |
| Married/domestic partnership | 165 (63) | 75 (58) |
| Divorced/separated/widowed | 31 (12) | 14 (11) |
| Never married | 65 (25) | 39 (30) |
| Missing | 1 (0) | 1 (1) |
| Inclusion Criteria |  |  |
| Central nervous system tumor<sup>a</sup> | 123 (47) | 56 (43) |
| Sarcoma<sup>b</sup> | 70 (27) | 41 (32) |
| Subsequent malignancy<sup>c</sup> | 69 (26) | 31 (24) |
| Missing | 0 (0) | 1 (1) |
| First-degree/second-degree relative with cancer | 204 (78) | 103 (80) |
| Mean No FDR/SDR with cancer (range) | 1.60 (0-8) | 1.58 (0-6) |
| No (%) of FDR/SDR with cancer |  |  |
| None | 58 (22.1) | 26 (20.2) |
| One | 89 (34.0) | 49 (38.0) |
| Two | 56 (21.4) | 25 (19.4) |
| Three | 34 (13.0) | 19 (14.0) |
| Four | 15 (5.7) | 6 (4.7) |
| Five | 5 (1.9) | 3 (2.3) |
| Six or more | 5 (1.9) | 2 (1.5) |
| Have health insurance | 247 (94) | 125 (97) |
| In a high deductible plan* | 87 (33) | 45 (35) |
| Have a usual source of care | 229 (87) | 117 (91) |
| Locale classification |  |  |
| City | 64 (24) | 28 (22) |
| Suburban | 76 (29) | 37 (29) |
| Rural | 120 (46) | 62 (48) |
| Missing/not classified | 2 (1) | 2 (2) |
| Total Social Vulnerability Index |  |  |
| Weighted average (SD) | 0.4 (0.2) | 0.4 (0) |
| Range | 0.1 – 0.9 | 0.1 – 0.9 |
| Access to internet |  |  |
| Access daily (mobile) | 258 (99) | 125 (97) |
| Access daily (computer) | 244 (93) | 111 (86) |
| Home | 171 (65) | 82 (64) |
| Work | 158 (60) | 71 (55) |
| Health literacy |  |  |
| Score range: 0 – 12, Mean (SD) | 3.0 (2.3) | 2.8 (2.2) |
| General depression |  |  |
| Score range: 4 – 20, Mean (SD) | 7.0 (3.4) | 6.7 (3.7) |
| General Anxiety |  |  |
| Score range: 4 – 20, Mean (SD) | 7.8 (3.3) | 7.8 (3.4) |
| Cancer-specific distress |  |  |
| Score range: 0 – 40, Mean (SD) | 7.1 (8.9) | 6.5 (8.6) |
| Knowledge of genetic disease |  |  |
| Score range: 0 – 16, Mean (SD) | 8.3 (3.5) | 8.3 (3.3) |

\*High deductible definition: $\geq \$1400$ .
\*\*Weight average: based on the census tract overlap with the zip code land area to create zip code-level SVI statistics. High values indicate greater social vulnerability
<sup>a</sup> 138/179(77%) Glioma, 24/179 (13%) Medulloblastoma, 17/179 (9%) Ependymoma

### Uptake of genetic services between remote telehealth genetic services and usual care

At six months, 43% (113/262) of participants in the remote telehealth arms received genetic services compared to 15% (19/129) in the usual care arm (OR = 4·4, 95% CI 2·5-8·0, p<.0001). This included higher rates of both counseling (42%, 111/262, vs. 15%, 19/129, p<0·0001) and genetic testing (19%, 49/262, vs. 9%, 12/129, p=0·020). While the primary aim of the ENGAGE study is to compare remote services (combined videoconference and telephone) with usual care, there were no significant differences in the uptake of genetic services between the two remote telehealth service arms (phone: 45%, 61/135, uptake; videoconference: 41%, 52/127, p = 0·57). Ten percent (5/49) of the participants in the remote telehealth arms who completed genetic testing were found to have genetic mutations (two *CHEK2*, one *TP53*, one *NF1*, and one *BARD1*), as compared to 8% (1/12) participants who completed genetic testing in the usual care arm, which was relevant to heterozygous carrier status for an autosomal recessive condition (Table 2).

**Table 2.**
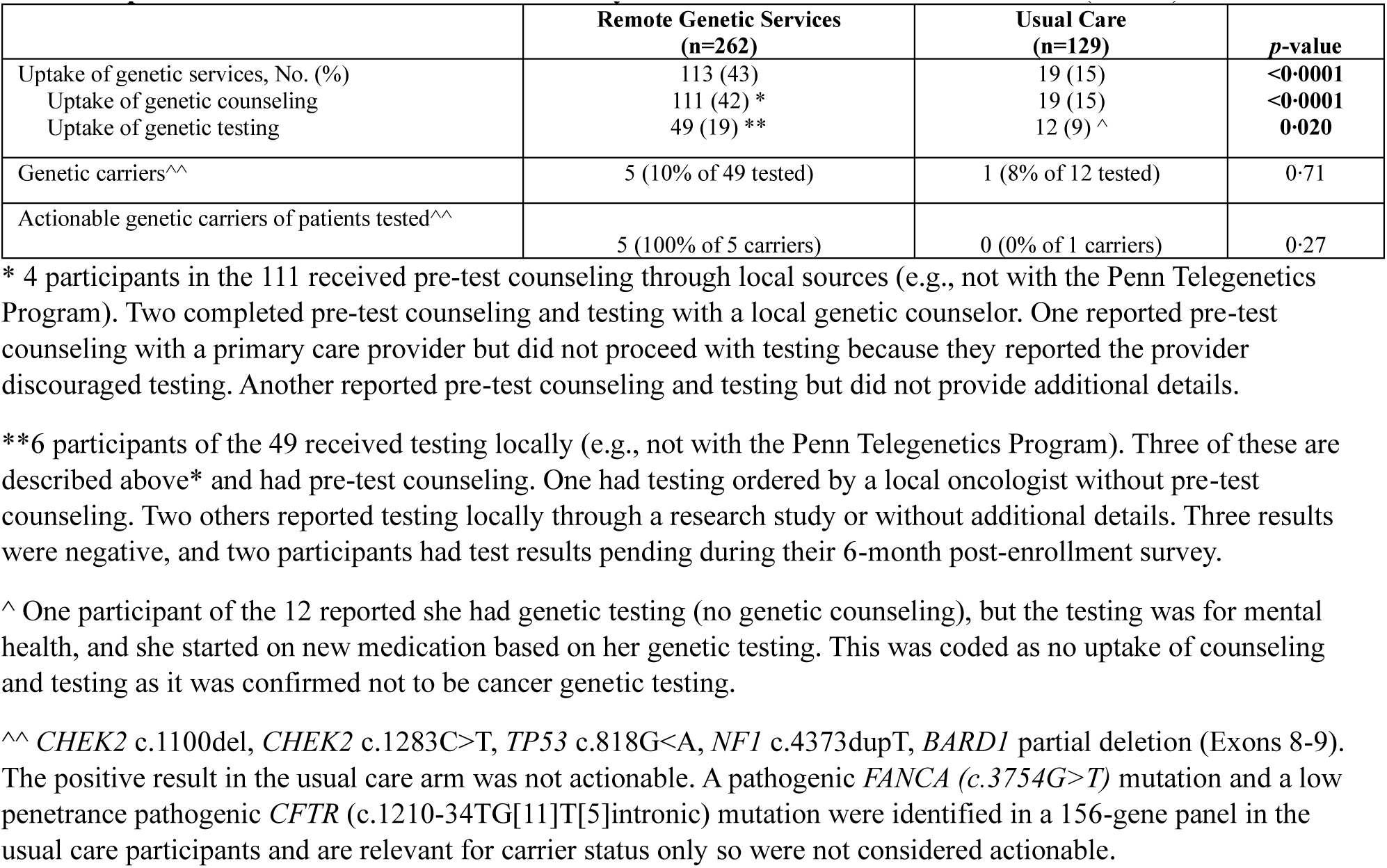
Uptake of Genetic Services at 6 months by intention-to-treat randomized arms (N=391).

Of the 12 participants in the usual care arm who reported having genetic counseling and testing, 75% (9/12) completed pretest counseling and testing with a local GC, and 25% (3/12) did so with their PCPs or other local healthcare providers. Seven participants received genetic counseling but did not proceed with testing. Among eight participants who shared their genetic test reports, testing included gene panels ranging from 32 to 156 genes, ordered through Invitae or Myriad Laboratories. Thirty-three percent (4/12) of participants reported having to pay out-of-pocket costs for their genetic testing, with unknown amounts.

In the remote telehealth arms, 56% (62/111) of participants did not complete testing after the counseling session. Genetic testing in the remote telehealth arms included panels for sarcoma (n = 18), CNS cancers (n = 15), multi-cancer (n = 5), hematologic malignancies (n = 5), and custom panels (n = 2). Some panels had additional genes based on personal or family history, with all tests ordered through Invitae or Ambry Laboratory. Forty-two percent (19/45) in the remote arms and 25% (1/4) in usual care reported out-of-pocket costs for their genetic testing (p=0.91); 45%, 9/20, paid < $200, 45%, 9/20 paid $200-250, 5%, 1/20, paid > $250, and 5%, 1/20, with unknown amount.

### Baseline factors associated with uptake of services

Secondary analyses explored baseline factors associated with the uptake of genetic services among all participants (n = 391), as well as factors associated with completing pretest counseling and genetic testing within the remote telehealth arms (n = 262). Overall, not having a high deductible health insurance plan (OR=1·67, 95% CI 1·00-2·81, p=0·049), higher perceived risk of cancer (OR=1·29, 95% CI 1·01-1·66, p=0·038), and having higher educational attainment were associated with greater uptake of genetic services (OR=1·54. 95% CI 1·09-2·20, p=0·012) Participants with a positive attitude toward genetic testing (OR=1·09, 95% CI 1·04-1·15, p=0·0003), higher genetic knowledge (OR=1·09/point, 95% CI 1·02-1·16, p=0·0075), and those who perceived the process as uncomplicated (OR=1·31, 95% CI 1·05-1·64, p=0·017) or not costing too much money (OR=1·51, 95% CI 1·17-1·96, p=0·0014) were also more likely to have genetic services. Conversely, participants experiencing higher levels of depression (OR= 0·91/point, 95% CI 0·85-0·98, p=0·0067) or anxiety (OR= 0·93/point, 95% CI 0·87-1·00, p=0·036) were less likely to complete genetic services (Figure 3a, Supplementary Table 2).

**FIGURES 3A and 3B.**
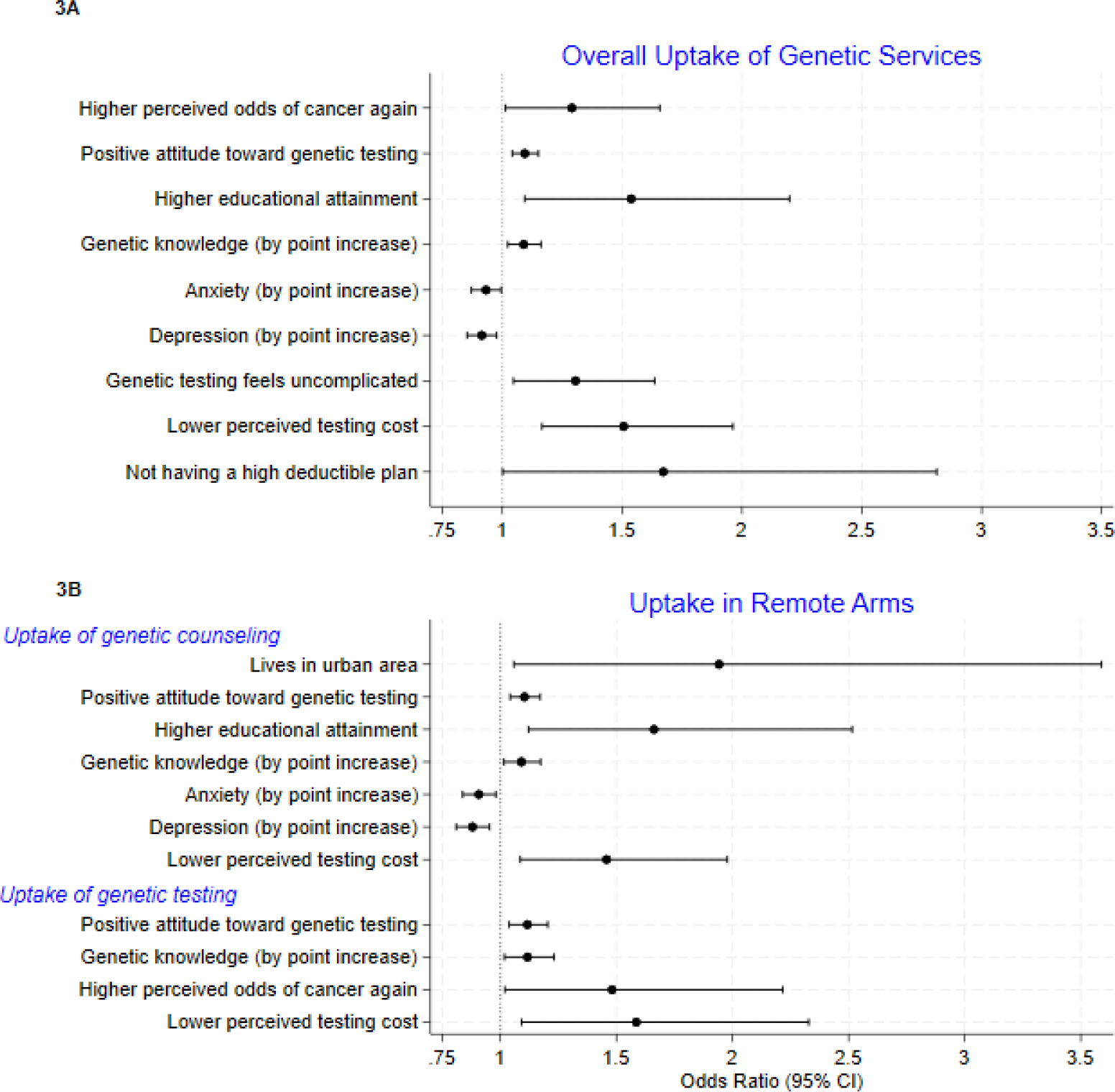
Factors associated with overall uptake of genetic services (N=391), and uptake of genetic counseling and genetic testing in the remote services arm (N=262).

Among the 262 participants in the remote telehealth arms, urban residence (OR=1·94, 95% CI 1·06-3·59, p=0·031), higher educational attainment (OR=1·66, 95% CI 1·12-2·52, p=0·010), a positive attitude toward genetic testing (OR=1·10, 95% CI 1·04-1·17, *p*=0·00067), lower perceived cost of testing (OR=1·46, 95% CI 1·09-1·98, p=0·011), higher genetic knowledge (OR=1·09, 95% CI 1·02-1·17, p=0·017) and lower levels of depression (OR=0·88, 95% CI 0·81-0·95, p=0·0013) and anxiety (OR=0·91, 95% CI 0·84-0·98, p=0·013) were significant predictors of genetic counseling. Factors associated with completing genetic testing included a higher perceived risk of cancer (OR=1·48, 95% CI 1·02-2·22, p=0·038), higher genetic knowledge (OR=1·12, 95% CI 1·02-1·23, p=0·019), a more positive attitude toward genetic testing (OR=1·12, 95% CI 1·04-1·20,p=0·0028), and a lower perceived cost of testing (OR= 1·59, 95% CI 1·09-2·33, p=0·014) (Figure 3b, Supplementary Table 3 and SupplementaryTable 4). Cancer type was not associated with uptake of counseling or testing in any of the above analyses.

### Qualitative data regarding barriers to genetic services

Among participants who completed the 6-month post-enrollment survey and reported not completing genetic services (n=160), the most cited reasons were time constraints (28%, 45/160), insufficient information about the process or rationale for testing (20%, 32/160), low perceived value (18%, 29/160) and concerns about cost or insurance coverage of testing (16%, 26/160) (Supplementary Table 5). Additional reasons included poor follow-through, access barriers, low prioritization, and lack of support from their provider. Most participants reported they did not discuss genetic testing with a local provider (70%, 53/86, in usual care; 90%, 98/108, in remote telehealth services) in response to a closed-ended (yes/no) question.

### Assessment of the in-home collaborative PCP model

A total of 84 local providers were registered with the Penn Telegenetics Program, which includes some patients in the waitlist arm. Of these, 52% (43/84) were male, 88% (76/84) were physicians, with the remainder being advanced practice providers, and 93% (82/84) specialized in internal or family medicine. Additionally, 77% (65/84) practiced in group settings, 19% (17/84) were in small practices, one worked in an academic practice, and the practice setting for the remainder was unknown.

For most patients (85%, 72/85), their physician registered in a timely manner allowing testing if they were interested. In six cases (7%, 6/85), registration delays led to testing delays, ranging from 15 to 63 days. In seven cases (8%, 7/85), local providers refused to register. In one of these cases, a second local provider was registered, but the participant could not be reached to schedule services.

## DISCUSSION

Identifying adult survivors of childhood cancer at risk for cancer predisposition syndromes is critical to optimize survivorship care and can also benefit their relatives and optimize their cancer prevention. To our knowledge, this is the first national randomized trial to investigate the effectiveness of remote telehealth genetic services in enhancing access to and uptake of genetic services among childhood cancer survivors as compared to usual care options in the survivors’ area. Many of these long-term childhood cancer survivors were diagnosed before genetic testing became clinically available, and no longer receive care at cancer centers, where genetic services are more accessible. Adult survivors of childhood cancer in this study who had access to remote telehealth genetic services were four times more likely to use genetic services than those in usual care. The low utilization rate (15%, 19/129) in the usual care arm was comparable to findings from a registry trial among childhood and young adult cancer survivors at a U.S. cancer center.^25^ Notably, 10% (5/49) of those who completed genetic testing in the remote telehealth arms had actionable results, enabling more personalized and precise survivorship care. For example, individuals with inherited mutations in *TP53* face a very high lifetime risk for multiple cancers, especially female breast cancer, which can happen at a young age. The NCCN has established guidelines for managing cancer risk in patients with these mutations.^26^

Despite remote access to telehealth genetic services being superior to usual care for uptake of genetic services, the proportion of survivors in the remote telehealth services arm completing pre-test counseling remained low at 43% (113/262), and fewer than 45% (49/111) of those who received genetic counseling pursued genetic testing. While easier access to genetic specialists through remote telehealth delivery significantly improves adherence to recommendations, barriers that hinder optimal use remain. Barriers like negative attitudes, low educational attainment, mental health comorbidities (e.g. depression and anxiety), and concerns about testing costs, align with findings from prior studies.^27^ Addressing these issues may require enhancing patient education about genetic services, providing additional reminders and support, and advocating for policy changes to lower the costs of testing and telehealth services. Equally important, integrating mental health support for co-morbid depression and anxiety, which are common in childhood cancer survivors,^28^ may be critical to ensuring uptake of genetic testing and optimal preventive care. Regardless, any incremental increase in testing uptake increases the number of genetic carriers identified. This can improve early detection and potentially reduce cancer incidence not only in childhood cancer survivors, but also in the family, and therefore has a potential incremental benefit.

Many childhood cancer survivors remain unaware of their risk for late effects, such as SMN. This is particularly true for those who completed treatment years ago and lack knowledge of their treatment history. ^29^ Without awareness of their SMN risk, survivors are less likely to recognize the value of genetic counseling and testing, and may not pursue appropriate genetic screening. Effective risk communication strategies during outreach and counseling are crucial to raising awareness. Additionally, just a fraction of those who received genetic counseling proceeded with genetic testing. Enhancing motivation for testing may require personalized decision aids, further education about its benefits, and financial support mechanisms to reduce concerns about testing costs. In this study, remote telehealth genetic counseling was provided free of charge to participants and funded by the study. However, costs for genetic testing, which are higher and more concerning to participants, were billed to their insurance, and some participants in both arms had out-of-pocket costs. Policy reforms that support mandating coverage of genetic testing, reducing out-of-pocket costs for testing, or providing subsidies for underinsured populations could increase the uptake of these services. Furthermore, these data demonstrate the value of providing centralized remote telehealth services for genetic medicine and advocating for policy reforms to cover telehealth services, ensuring that all patients, including those in rural and more remote locations, have access to critical services that can’t be adequately provided in their usual and local settings. At the time the study was conducted, there were no clear pathways for coverage of remote genetic counseling, but these have evolved and are currently being considered by the Penn Telegenetics Program, as long as telehealth visits in general continue to be covered by the Centers for Medicare & Medicaid Services (CMS). This is especially critical given the adversities faced by many childhood cancer survivors, such as lower educational attainment, unemployment or low-skilled jobs, inadequate health insurance, and limited income ^30^

In this study, most healthcare providers were willing to collaborate with remote GCs to order and manage genetic testing for their high-risk patients. Studies have shown that primary care providers are not always aware of COG guidelines in managing their childhood cancer survivors; however, they are willing to care for these survivors.^11^ The high willingness of local providers to collaborate on testing demonstrates strong potential for widespread adoption of the In-Home Collaborative PCP Model, underscoring the importance of facilitating provider engagement in survivorship care and care coordination.

As genetic testing has become more efficient and affordable, expanding access through approaches, such as remote telehealth genetic services, could make broader testing feasible for all childhood cancer survivors. With 50% of genetic counselors reporting that they practice in academic medical centers, cancer centers can provide critical access to genetic services. However, families may be overwhelmed by their diagnosis and treatment, concerned about cost and insurance coverage, or not offered testing without clear hereditary indicators, which can limit uptake. Moreover, many newly diagnosed adolescents and young adults (AYAs) with cancer are treated in community settings, also limiting access to genetic services. Integrating genetic assessment into survivorship care (the majority of which is delivered in the community), supplemented by innovative delivery models, such as remote telehealth genetic services, can be an effective approach to ensuring that all survivors have access to genetic risk assessment.

This study has several strengths, including being the largest randomized trial studying remote genetic services in childhood cancer survivors, using a “real-world” approach and providing services in the home, and enrolling a racially, socioeconomically, and geographically diverse group of childhood cancer survivors. There were also limitations that should be considered when interpreting the results. The ultimate racial and ethnic makeup of recruited patients may not be generalizable to the US population. The study was conducted during the COVID-19 pandemic, and preventive care, such as genetic counseling and testing, may have been a lower priority. A few primary care providers declined to collaborate, preventing remote testing for a few patients. Of note, this study was conducted prior to the release of the COG Long-Term Follow-Up guidelines for genetic testing.^9^ These guidelines can potentially increase awareness among survivors and providers, as well as support for insurance coverage. Furthermore, additional childhood cancer survivors may now be candidates for testing beyond the narrower inclusion criteria in this study. The release of these guidelines may help address some barriers and provide even stronger support for offering remote services, thereby increasing the uptake of guideline-based survivorship care for these survivors.

## CONCLUSION

Remote centralized telehealth genetic services show significant promise in enhancing access to and adherence to survivorship care recommendations. To maximize their impact, we must address the remaining barriers through targeted interventions, such as personalized risk communication, policy reform, and sustainable models like the PCP collaboration. By addressing these gaps, we can advance survivorship care, improve long-term outcomes, and ensure equitable access for childhood cancer survivors and their families.

## Data Availability

All data produced in the present study are available upon reasonable request to the authors.

https://viz.stjude.cloud/community/cancer-survivorship-community~4/publications.

## Contributors

TOH, ARB, BE, LGF, EBE, and MP designed the study. MAA, RM, DO, BM, and HG provided administrative support and assisted with data collection and participant recruitment. SH, EMW, CNC, and SB provided genetic counseling and assisted in data collection. TOH, ARB, and BE developed a data analysis plan and conducted data analysis. TOH, ARB, BE, KCO, KRK, and GTA accessed and verified the raw data used in the study. All authors contributed to the data interpretation and writing and approved the final manuscript for publication.

## Data Sharing Statement

The Childhood Cancer Survivor Study is a US National Cancer Institute-funded resource (U24 CA55727) to promote and facilitate research among long-term survivors of cancer diagnosed during childhood and adolescence. CCSS data are publicly available on dbGaP at https://www.ncbi.nlm.nih.gov/gap/ through its accession number phs001327.v2.p1. and on the St Jude Survivorship Portal within the St. Jude Cloud at https://survivorship.stjude.cloud/. In addition, utilization of the CCSS data that leverages the expertise of CCSS Statistical and Survivorship research and resources will be considered case-by-case. For this utilization, a research Application of Intent followed by an Analysis Concept Proposal must be submitted for evaluation by the CCSS Publications Committee. Users interested in utilizing this resource are encouraged to visit http://ccss.stjude.org. Full analytical data sets associated with CCSS publications since January of 2023 are also available on the St. Jude Survivorship Portal at https://viz.stjude.cloud/community/cancer-survivorship-community∼4/publications.

