## Supplementary Table 1 for "The ENGAGE Study: A Randomized Trial Optimizing Uptake of Germline Cancer Genetic Services in Childhood Cancer Survivors"

**Supplementary Materials**

| Table of Contents | Page |
|---|---|
| Supplementary Table 1. Demographics of Responders versus Non-Responders. | 2 |
| Supplementary Table 2a. Unadjusted odds ratios for factors associated with uptake of genetic services in both arms (N = 391). | 3 |
| Supplementary Table 2b. Multivariable adjusted model for factors associated with uptake of genetic services in both arms. | 4 |
| Supplementary Table 3a. Unadjusted odds ratios for factors associated with pre-test genetic counseling in the remote arm (n = 262). | 5 |
| Supplementary Table 3b. Multivariable adjusted model for factors associated with pre-test genetic counseling in the remote arm. | 6 |
| Supplementary Table 4a. Unadjusted odds ratios for factors associated with genetic testing uptake in the remote arm (n = 262). | 7 |
| Supplementary Table 4b. Multivariable adjusted model for factors associated with genetic testing in the remote arm. | 8 |
| Supplementary Table 5. Patients reported barriers to genetic services in both arms (n = 160). | 9 |
| Supplementary Table 6. List of subsequent malignancies by tumor types | 10 |

**Supplementary Table 1. Demographics of Responders versus Non-Responders.**

| | Consented (N = 391) | Refused/No Baseline Survey/ineligible post-consented (N = 164) | *p* - value |
|---|---|---|---|
| **Age at Invite** | | | 0·054 |
| Mean (SD) | 43·97 (9·66) | 45·86 (10·96) | |
| Median (Q1, Q3) | 43·0 (37·0, 51·0) | 46·0 (37·0, 55·0) | |
| Min, Max | 24·0, 71·0 | 23·0, 69·0 | |
| **Sex·** | | | 0·66 |
| Male | 181 (46·3%) | 80 (48·8%) | |
| Female | 210 (53·7%) | 84 (51·2%) | |
| **Race** | | | 0·12 |
| White | 313 (80·1%) | 122 (74·4%) | |
| Black | 43 (11·0%) | 31 (18·9%) | |
| American Indian/Alaskan Native | 3 (0·8%) | 0 (0·0%) | |
| Asian or Pacific Islander | 18 (4·6%) | 5 (3·0%) | |
| Other | 12 (3·1%) | 4 (2·4%) | |
| Unknown | 2 (0·5%) | 2 (1·2%) | |
| **Hispanic** | | | 0·77 |
| Yes | 17 (4·3%) | 5 (3·0%) | |
| No | 360 (92·1%) | 153 (93·3%) | |
| Unknown | 14 (3·6%) | 6 (3·7%) | |
| **ENGAGE refusal reason** | | | . |
| Not Comfortable with Technology | 0 (0·0%) | 8 (4·9%) | |
| Not Interested | 0 (0·0%) | 61 (37·2%) | |
| Other | 0 (0·0%) | 53 (32·3%) | |
| Privacy/Security Concerns | 0 (0·0%) | 6 (3·7%) | |
| Too Busy | 0 (0·0%) | 12 (7·3%) | |
| Did not complete the baseline survey | 0 (0·0%) | 20 (12·2%) | |
| Missing information | 0 (0·0%) | 4 (2·4%) | |

**Supplementary Table 2a. Unadjusted odds ratios for factors associated with uptake of genetic services in both arms (N = 391). Two Likert measures were reverse coded to ease interpretation.**

| | | **Did not have genetic services** | **Did have genetic services** | | |
|---|---|---|---|---|---|
| **Factor*** | N | Mean (SD) or N (%) | Mean (SD) or N (%) | Unadjusted Odds Ratio (95%CI) | *p*-value |
| Higher perceived odds of getting cancer again (per ordinal Likert increase) | 389 | 3.65 (0.96) | 3.86 (0.86) | 1.29 (1.01, 1.66) | 0.038 |
| Positive attitude toward genetic testing (per point increase) | 390 | 29.50 (4.32) | 31.20 (4.47) | 1.09 (1.04, 1.15) | 0.00030 |
| Higher education attainment (per ordinal increase) | 390 | No college: 32 (12.4%)<br>Some college/Trade school: 74 (28.7%)<br>College+: 152 (58.9%) | 10 (7.6%)<br>26 (19.7%)<br>96 (72.7%) | 1.54 (1.09, 2.20) | 0.012 |
| Genetic knowledge (per point increase in knowledge) | 390 | 7.96 (3.49) | 8.94 (3.21) | 1.09 (1.02, 1.16) | 0.0075 |
| Anxiety (per point increase in anxiety) | 387 | 8.07 (3.47) | 7.31 (3.02) | 0.93 (0.87, 1.00) | 0.036 |
| Depression (per point increase in depression) | 388 | 7.28 (3.77) | 6.28 (2.78) | 0.91 (0.85, 0.98) | 0.0067 |
| Getting genetic testing feels complicated (per Likert increase) | 390 | 2.61 (0.96) | 2.36 (1.02) | Reverse coded OR<br>1.31 (1.05, 1.64) | 0.017 |
| Genetic testing will cost me too much money (per ordinal Likert increase) | 390 | 3.15 (0.84) | 2.85 (0.90) | Reverse coded OR<br>1.51 (1.17, 1.96) | 0.0014 |
| Not having a high-deductible plan (relative to having one) | 372 | Deductible at least $1400: 92 (37.6%)<br>Deductible <$1400: 92 (37.6%)<br>Don't know: 61 (24.9%) | 40 (31.5%)<br>67 (52.8%)<br>20 (15.7%) | 1.67 (1.00, 2.81) | 0.0491 |

*No factors were statistically significant in the multivariable model.

SD=Standard deviation, N=number of participants

**Supplementary Table 2b. Multivariable adjusted odds ratios for factors associated with GC or GT uptake (all consented). N=364**

| Any genetic services | Odds ratio | Std. err. | z | P>\|z\| | [95% conf. interval] | |
|---|---|---|---|---|---|---|
| **Randomization Arm** | | | | | | |
| Video/Phone | 4.53 | 1.35 | 5.05 | 0.000 | 2.52 | 8.14 |
| Usual care | 1.00 | (base) | | | | |
| **Genetic Knowledge Score** | 1.06 | 0.04 | 1.44 | 0.15 | 0.98 | 1.14 |
| **PROMIS Depression** | 0.93 | 0.05 | -1.25 | 0.21 | 0.84 | 1.04 |
| **Perceived odds of getting cancer** | 1.32 | 0.19 | 1.96 | 0.050 | 1.00 | 1.75 |
| **Number FDRs/SDRs with cancer** | 1.02 | 0.09 | 0.17 | 0.86 | 0.85 | 1.21 |
| **Urban Location** | | | | | | |
| Rural/Suburban | 1.00 | (base) | | | | |
| Urban | 1.46 | 0.42 | 1.32 | 0.19 | 0.83 | 2.56 |
| **Attitudes to genetic testing** | 1.05 | 0.03 | 1.57 | 0.12 | 0.99 | 1.12 |
| **High deductible plan** | | | | | | |
| Yes | 1.00 | (base) | | | | |
| No | 1.58 | 0.44 | 1.64 | 0.10 | 0.91 | 2.75 |
| Don’t know | 0.82 | 0.30 | -0.52 | 0.60 | 0.40 | 1.70 |
| **Testing feels complicated (reversed)** | 1.04 | 0.15 | 0.25 | 0.80 | 0.78 | 1.38 |
| **Cost concerns (reversed)** | 1.17 | 0.19 | 0.98 | 0.33 | 0.85 | 1.62 |
| **PROMIS Anxiety** | 0.98 | 0.05 | -0.42 | 0.67 | 0.88 | 1.09 |
| **Ordinal education** | 1.31 | 0.25 | 1.39 | 0.17 | 0.90 | 1.90 |
| **Age** | 1.00 | 0.01 | -0.34 | 0.73 | 0.97 | 1.02 |
| **Self-efficacy** | 0.96 | 0.04 | -0.95 | 0.34 | 0.89 | 1.04 |
| **Sex** | | | | | | |
| Male | 1.00 | (base) | | | | |
| Female | 0.93 | 0.24 | -0.30 | 0.77 | 0.56 | 1.53 |
| Intercept | 0.01 | 0.01 | -3.17 | 0.002 | 0.00 | 0.17 |

**Supplementary Table 3a. Unadjusted odds ratios for factors associated with pre-test genetic counseling in the remote arm (n = 262).**

| | | **Did not have pre-test genetic counseling** | **Did have pre-test genetic counseling** | | |
|---|---|---|---|---|---|
| **Factor*** | N | Mean (SD) or N (%) | Mean (SD) or N (%) | Unadjusted Odds Ratio (95%CI) | *p*-value |
| Positive attitude toward genetic testing (per point increase) | 261 | 29.23 (4.24) | 31.11 (4.49) | 1.10 (1.04, 1.17) | **0.00067** |
| Higher education attainment (per ordinal increase) | 262 | No College: 20 (13.2%)<br>Some College/Trade school: 42 (27.8%)<br>Collage+" 89 (58.9%) | 10 (9.0%)<br>15 (13.5%)<br>86 (77.5%) | 1.66 (1.12, 2.52) | **0.010** |
| Genetic knowledge (per point increase in knowledge) | 261 | 7.86 (3.68) | 8.90 (3.16) | 1.09 (1.02, 1.17) | **0.017** |
| Anxiety (per point increase in anxiety) | 259 | 8.28 (3.53) | 7.24 (2.96) | 0.91 (0.84, 0.98) | **0.013** |
| Depression (per point increase in depression) | 259 | 7.60 (3.77) | 6.24 (2.72) | 0.88 (0.81, 0.95) | **0.0013** |
| Lower perceived cost of testing (per ordinal Likert increase) | 261 | 3.13 (0.85) | 2.84 (0.91) | Reverse Coded<br>1.46 (1.09, 1.98) | **0.011** |
| Live in an urban area (vs. rural/suburban combined) | 260 | 121 (80.7%)<br>29 (19.3%) | 75 (68.2%)<br>35 (31.8%) | 1.94 (1.06, 3.59) | **0.031** |

*No factors were statistically significant in the adjusted model.

SD=Standard deviation, N=number of participants

**Supplementary Table 3b. Multivariable adjusted odds ratios for factors associated with GC uptake (Remote arms only). N=243**

| Any genetic counseling (pre-test) | Odds ratio | Std. err. | z | P>\|z\| | [95% conf. interval] | |
|---|---|---|---|---|---|---|
| **Genetic Knowledge Score** | 1.06 | 0.05 | 1.35 | 0.18 | 0.97 | 1.16 |
| **PROMIS Depression** | 0.94 | 0.06 | -1.00 | 0.32 | 0.83 | 1.06 |
| **Perceived odds of getting cancer** | 1.22 | 0.19 | 1.26 | 0.21 | 0.90 | 1.67 |
| **Number FDRs/SDRs with cancer** | 0.99 | 0.10 | -0.07 | 0.94 | 0.81 | 1.22 |
| **Urban Location** | | | | | | |
| Rural/Suburban | 1.00 | (base) | | | | |
| Urban | 1.75 | 0.57 | 1.71 | 0.087 | 0.92 | 3.33 |
| **Attitudes to genetic testing** | 1.05 | 0.04 | 1.31 | 0.19 | 0.98 | 1.13 |
| **High deductible plan** | | | | | | |
| Yes | 1.00 | (base) | | | | |
| No | 1.65 | 0.53 | 1.55 | 0.12 | 0.88 | 3.09 |
| Dont_know | 1.12 | 0.46 | 0.28 | 0.78 | 0.50 | 2.49 |
| **Testing feels complicated (reversed)** | 1.08 | 0.19 | 0.45 | 0.66 | 0.77 | 1.52 |
| **Cost concerns (reversed)** | 1.06 | 0.20 | 0.30 | 0.76 | 0.73 | 1.54 |
| **PROMIS Anxiety** | 0.96 | 0.06 | -0.71 | 0.48 | 0.85 | 1.08 |
| **Ordinal education** | 1.45 | 0.32 | 1.66 | 0.097 | 0.94 | 2.24 |
| **Age** | 1.00 | 0.02 | -0.13 | 0.90 | 0.97 | 1.03 |
| **Self-efficacy** | 0.99 | 0.05 | -0.16 | 0.88 | 0.91 | 1.09 |
| **Sex** | | | | | | |
| Male | 1.00 | (base) | | | | |
| Female | 0.87 | 0.26 | -0.48 | 0.63 | 0.49 | 1.55 |
| Intercept | 0.03 | 0.06 | -2.00 | 0.046 | 0.00 | 0.94 |

**Supplementary Table 4a. Unadjusted odds ratios for factors associated with genetic testing uptake in the remote arm (n = 262).**

| | | **Did not have genetic testing** | **Had genetic testing** | | |
|---|---|---|---|---|---|
| **Factor*** | N | Mean (SD) | Mean (SD) | Unadjusted Odds Ratio (95%CI) | *p*-value |
| Positive attitude toward genetic testing (per point increase) | 261 | 29.64 (4.37) | 31.73 (4.38) | 1.12 (1.04, 1.20) | **0.0028** |
| Genetic knowledge (per point increase in knowledge) | 261 | 8.06 (3.57) | 9.35 (3.02) | 1.12 (1.02, 1.23) | **0.019** |
| Lower perceived cost of testing (per ordinal Likert increase) | 261 | 3.07 (0.88) | 2.71 (0.87) | Reverse Coded<br>1.59 (1.09, 2.33) | **0.014** |
| Higher perceived odds of getting cancer again (per ordinal Likert increase) | 261 | 3.64 (0.99) | 3.96 (0.76) | 1.48 (1.02, 2.22) | **0.038** |

*No factors were statistically significant in the adjusted model.

SD=Standard deviation, N=number of participants

**Supplementary Table 4b. Multivariable adjusted odds ratios for factors associated with GT uptake (Remote arms). N=243**

| Any genetic testing | Odds ratio | Std. err. | z | P>\|z\| | [95% conf. interval] | |
|---|---|---|---|---|---|---|
| **Genetic Knowledge Score** | 1.09 | 0.06 | 1.49 | 0.14 | 0.97 | 1.22 |
| **PROMIS Depression** | 1.00 | 0.08 | -0.02 | 0.98 | 0.86 | 1.17 |
| **Perceived odds of getting cancer** | 1.56 | 0.35 | 1.96 | 0.050 | 1.00 | 2.42 |
| **Number FDRs/SDRs with cancer** | 0.93 | 0.13 | -0.56 | 0.58 | 0.71 | 1.21 |
| **Urban Location** | | | | | | |
| Rural/Suburban | 1.00 | (base) | | | | |
| Urban | 1.60 | 0.63 | 1.20 | 0.23 | 0.74 | 3.48 |
| **Attitudes to genetic testing** | 1.09 | 0.05 | 1.80 | 0.072 | 0.99 | 1.19 |
| **High-deductible plan** | | | | | | |
| Yes | 1.00 | (base) | | | | |
| No | 1.35 | 0.55 | 0.75 | 0.45 | 0.61 | 2.99 |
| Dont_know | 1.02 | 0.55 | 0.04 | 0.97 | 0.35 | 2.96 |
| **Testing feels complicated (reversed)** | 1.02 | 0.22 | 0.08 | 0.93 | 0.66 | 1.56 |
| **Cost concerns (reversed)** | 1.34 | 0.31 | 1.26 | 0.22 | 0.85 | 2.11 |
| **PROMIS Anxiety** | 0.93 | 0.07 | -0.89 | 0.38 | 0.80 | 1.09 |
| **Ordinal education** | 0.85 | 0.23 | -0.62 | 0.54 | 0.50 | 1.44 |
| **Age** | 0.98 | 0.02 | -1.15 | 0.25 | 0.94 | 1.02 |
| **Self-efficacy** | 1.01 | 0.06 | 0.21 | 0.83 | 0.90 | 1.13 |
| **Sex** | | | | | | |
| Male | 1.00 | (base) | | | | |
| Female | 0.76 | 0.28 | -0.76 | 0.45 | 0.37 | 1.55 |
| Intercept | 0.00 | 0.01 | -2.70 | 0.007 | 0.00 | 0.20 |

**Supplementary Table S5. Patients reported barriers to genetic services in both arms (n = 160).**

| Coded themes and examples of patient quotes | Overall (N=160) | Remote Genetic Services (n=90) | Usual Care (n=70) |
|---|---|---|---|
| **Time constraints, No. (%)**<br>"*I've been busy working full time for the first time."*<br>*"Busy with family."*<br>*"Could not get around to it, but will attempt to in the near future."* | 45 (28·1) | 22 (24·0) | 23 (32·9) |
| **Not enough information regarding the process or rationale, No. (%)**<br>*"Wasn't clear on how, who, and where to do this. It is too difficult for me to understand and process, possibly due to my chemo brain."*<br>*"I don't know. That's not something I've ever been advised to do."*<br>*"Didn't know what they were talking about."*<br>*Confusion about the process itself primarily, cost secondary."* | 32 (20%) | 18 (20·0) | 14 (18·7) |
| **Low perceived value, no interest, No. (%)**<br>*"Don't feel I need it."*<br>*"Diagnosis is already confirmed. No need for genetic counseling at this time."*<br>*"Not necessary, as I don't have any health issues."* | 29 (18·1) | 16 (17·8) | 13 (18·6) |
| **Concerns about cost or insurance coverage, No. (%)**<br>"*I also cannot afford to pay for anything right now. I have no extra funds to spend on testing/doctor appointments. Unfortunately, we're using every penny we've got just to get by."*<br>*"I don't think it would be affordable. Insurance companies are reluctant to cover anything these days."*<br>*"I also had to do all the leg work at the expense of the testing, call my insurance company, and work with the clinic. It took about half a day to figure out if it would be covered. Even as I went for my appointment, I wasn't 100% sure if the test itself would be covered. I feel like this was a stressor."* | 26 (16·3) | 10 (9%) | 16 (22·8) |
| **Poor follow-through or difficult scheduling, No. (%)**<br>*"Thought they were supposed to call me automatically.:*<br>*"I forgot to schedule."* | 18 (11·3) | 18 (20) | 0 |
| **Hard to access, No. (%)**<br>"*Small town…I don't know how or where to receive testing."*<br>*"No computer at home."*<br>*"I reached out to my provider (they don't do that) and a place I found online without response."*<br>*"I don't drive and live far from the hospital to get it done. Also, my family doctor said they do not do it there, so that was not an option. I was told I would need to go see a specialist, and I just cannot do that. It was too difficult to get done."* | 13 (8·1) | 5 (5·5) | 8 (10·7) |
| **Not a priority, No. (%)**<br>*"I was in the middle of a job search, and it was not a priority."*<br>*"One of my children was diagnosed with Autism, and I have been super busy trying to get things situated with him."*<br>*"Have let myself become distracted with other responsibilities and have ignored my personal health care interests."* | 7 (4·4) | 7 (7·7) | 0 |
| **Provider didn't support, No. (%)**<br>*"I asked my provider, but they deemed it unnecessary and could not provide a referral or recommendation."* | 4 (2·5) | 0 | 4 (5·7) |

Other less frequently reported barriers were anxiety (n=1 in usual care), discomfort in sharing personal information (n=1 in remote services), and planning to test later (n=2 in remote services). Some said they might test in the future.

**Supplementary Table 6. List of subsequent malignancies by tumor types**

| | SMN 1 | SMN 2 |
|---|---|---|
| Glioma (n=4) | Papillary adenocarcinoma (Thyroid gland); Astrocytoma (Brain, unspecific); Glioblastoma (Temporal lobe); Oligodendroglioma (Overlapping lesion of brain) | |
| Ependymoma (n=2) | Papillary microcarcinoma (Thyroid gland); Papillary carcinoma columnar cell (Thyroid gland) | |
| Medulloblastoma (n=1) | Papillary adenocarcinoma (Thyroid gland) | |
| Osteosarcoma (n=5) | Adenocarcinoma (Parotid gland); Adenocarcinoma (Rectosigmoid junction); Papillary adenocarcinoma (Thyroid gland); Kaposi sarcoma (Skin of trunk); Chronic eosinophilic leukemia | |
| Rhabdomyosarcoma (n=1) | Papillary adenocarcinoma (Thyroid gland) | |
| Non-Rhabdomyosarcoma (n=2) | Papillary adenocarcinoma (Thyroid gland); Hodgkin lymphoma (Lymph node, unspecific) | |
| Ewing Sarcoma (n=2) | Adenocarcinoma (Prostate); Undifferentiated sarcoma (Long bones of lower limb) | |
| Sarcoma, NOS (n=0) | | |
| Leukemia (n=24) | Adenocarcinoma (Appendix); Carcinoid tumor (Ileum); 2 Papillary adenocarcinoma (Thyroid gland); 3 Follicular carcinoma (Thyroid gland); Papillary microcarcinoma (Thyroid gland); 2 **Endometrioid adenocarcinoma (Endometrium)**; Mucoepidermoid carcinoma (Parotid gland); Mucinous adenocarcinoma (Endometrium); Infiltrating duct carcinoma (Breast); Lobular carcinoma (Breast); **Acinar cell carcinoma (Parotid gland)**; Malignant melanoma (Skin of trunk); Superficial spreading melanoma (Skin of lower limb); Leiomyosarcoma (connective & soft tissue lower limb); Astrocytoma (Frontal lobe); Astrocytoma (Temporal lobe); Hodgkin lymphoma (Lymph node, unspecific); Malignant lymphoma, large B-cell diffuse (Nasopharynx, unspecific); Chronic myeloid leukemia, Melanoma (Skin of scalp and neck) | **Papillary adenocarcinoma (Thyroid gland)**; **Papillary carcinoma, follicular variant (Thyroid gland)** |
| Hodgkin's Disease (n=20) | Transitional cell carcinoma (Trigone of bladder); Adenocarcinoma (Lung); 2 Papillary adenocarcinoma (Thyroid gland); Oxyphilic adenocarcinoma (Thyroid gland); 3 Follicular adenocarcinoma (Thyroid gland); Papillary microcarcinoma (Thyroid gland); **Mucinous adenocarcinoma (Lung)**; 4 Infiltrating duct carcinoma (Breast); Malignant fibrous histiocytoma (Head, face); Ewing's sarcoma (Lower limb); **Hodgkin lymphoma (Inguinal & lower limb lymph node)**; Follicular lymphoma, grade 3 (Ileum); Marginal zone B-cell lymphoma (Intrathoracic lymph node); Malignant lymphoma, large B-cell, diffuse | **Malignant melanoma (Skin of trunk)**; **Malignant lymphoma, non-Hodgkin** |
| Kidney (n=9) | Transitional cell carcinoma (Bladder, unspecific); Adenocarcinoma(Sigmoid colon); Adenocarcinoma (Prostate); 3 Papillary adenocarcinomas (Thyroid gland); Endometrioid adenocarcinoma (Endometrium); Signet ring cell carcinoma (Pyloric antrum); Neurilemoma (Spinal cord) | |
| Neuroblastoma (n=1) | Infiltrating duct carcinoma (Breast) | |
| Non-Hodgkin's (n=3) | Papillary adenocarcinoma (Thyroid gland); Acinar cell carcinoma (Prostate); B-cell lymphocytic leukemia/small lymphocytic lymphoma | |
| Total | n = 74 | n = 4 |
